# Low CD4/CD8 ratio predicts cancer risk among adults with HIV

**DOI:** 10.1101/2021.07.21.21260588

**Authors:** Jessica L. Castilho, Aihua Bian, Cathy A. Jenkins, Bryan E. Shepherd, Keith Sigel, M. John Gill, Mari M. Kitahata, Michael J. Silverberg, Angel M. Mayor, Sally B. Coburn, Dorothy Wiley, Chad J. Achenbach, Vincent C. Marconi, Ronald J. Bosch, Michael A. Horberg, Charles Rabkin, Sonia Napravnik, Richard M. Novak, W. Christopher Mathews, Jennifer E. Thorne, Jing Sun, Keri N. Althoff, Richard D. Moore, Timothy R. Sterling, Staci L. Sudenga, the North American AIDS Cohort Collaboration on Research and Design (NA-ACCORD) of the International Epidemiology Databases to Evaluate AIDS (IeDEA)

## Abstract

**Background:** Independent of CD4 cell count, low CD4/CD8 ratio in people with HIV (PWH) is associated with deleterious immune senescence, activation, and inflammation, which may contribute to carcinogenesis and excess cancer risk. We examined whether low CD4/CD8 ratios predicted cancer among PWH in the USA and Canada.

**Methods:** We examined all cancer-free PWH with one or more CD4/CD8 values from NA-ACCORD observational cohorts with validated cancer diagnoses between 1998-2016. We evaluated the association between time-lagged CD4/CD8 ratio and risk of specific cancers in multivariable, time-updated Cox proportional hazard models using restricted cubic spines.

Models were adjusted for age, sex, race/ethnicity, hepatitis C virus, and time-updated CD4 cell count, HIV RNA, and history of AIDS-defining illness.

**Results:** Among 83,893 PWH, there were 5,628 incident cancers, including lung cancer (n=755), Kaposi sarcoma (KS, n=501), non-Hodgkin lymphoma (NHL, n=497), and anal cancer (n=439). Median age at cohort entry was 43 years, 87% were male, and 43% were white. Overall median six-month lagged CD4/CD8 ratio was 0.52 (interquartile range: 0.30-0.82). Compared with six-month lagged CD4/CD8=0.80, CD4/CD8=0.30 was associated with increased risk of any incident cancer (adjusted hazard ratio = 1.24 [95% confidence interval: 1.14-1.35]). CD4/CD8 ratio was also inversely associated with NHL, KS, lung cancer, anal cancer, and colorectal cancer in adjusted analyses (all *p*<0.05). Results were similar using 12-, 18-, and 24-month lagged CD4/CD8 values.

**Conclusions:** Low CD4/CD8 ratio up to 24 months prior to cancer diagnosis was independently associated with increased cancer risk in PWH and may serve as a clinical biomarker.

## Introduction

While people with HIV (PWH) are living longer with use of effective antiretroviral therapy (ART), they remain at an increased risk for cancer compared to the general U.S. population [1-5]. In addition to AIDS-defining cancers (ADCs) including Kaposi sarcoma, non-Hodgkin lymphomas (NHL) and cervical cancer, the risk of several non-AIDS-defining cancers (NADCs) such as liver, lung, anal, oral pharyngeal cancers and Hodgkin lymphoma remains increased in PWH [6-9]. Cancer mortality is also significantly higher among PWH compared to the general population [10, 11]. Approximately 10% of all deaths among PWH prescribed ART in the U.S. between 1995-2009 were attributed to cancer; with NHL, lung, and liver cancer contributing the highest number of deaths [10].

Increased cancer risk in PWH is attributed to HIV-induced immunodeficiency, immune dysfunction, coinfection with oncogenic viruses, and high prevalence of behavioral risk factors for cancer, including smoking [12-15]. Altered immunity, not simply immunodeficiency, is one proposed mechanism driving the excess risk of cancer in PWH. Decreased immune surveillance and increased cellular replication associated with immune senescence and activation, and inflammation increase carcinogenesis [16, 17]. The prevalence of oncogenic viruses, such as human papillomavirus (HPV) and hepatitis C virus (HCV), is high among PWH [15]. Lastly, 20% of all incident cancers among PWH is attributed to smoking [14].

In PWH, CD4 and CD8 measures are generally readily available clinical tests. Low CD4/CD8 ratio inversely correlates with T lymphocyte replicative senescence, activation and exhaustion, each a measure of immunosenescence and systemic inflammation [18-20]. Low CD4/CD8 ratio is also associated with older age, cytomegalovirus (CMV) coinfection, and low CD4 nadir in PWH, and has been associated with risk of non-AIDS outcomes [18, 21-27]. We sought to determine whether CD4/CD8 ratio, as a marker of immune dysregulation, was associated with cancer risk, with the hypothesis that CD4/CD8 ratio would be inversely associated with incident cancer diagnoses among PWH.

## Methods

This study examined incident cancer diagnoses and longitudinal CD4/CD8 data from the North American AIDS Cohort Collaboration on Research and Design (NA-ACCORD), the largest multi-site consortium of observational clinical cohorts of PWH in the United States and Canada [28]. Participating cohorts contributed patient-level demographic, behavioral, and clinical data including diagnoses, medications, and laboratory values to the central Data Management Center for quality assessment and harmonization. All cancer diagnoses in the NA-ACCORD were validated by standardized medical record documentation that included abstraction of histopathologic information or linkage with cancer registries [4]. The institutional review boards at participating cohorts, Johns Hopkins University, and Vanderbilt University have approved human subject activities conducted within the NA-ACCORD related to this study.

NA-ACCORD includes PWH ≥18 years of age with at least two clinic visits within the first year of cohort enrollment. A total of 22 observational cohorts in NA-ACCORD collected validated cancer diagnoses from January 1998-December 2016. This study excluded seven cohorts where ≥25% of PWH had no CD8 cell counts recorded. All PWH with any cancer diagnosis before or within the first six months of cohort entry or a cancer with an unknown diagnosis date were excluded. Only PWH with a minimum of 12 months of clinic follow-up were included in the primary analysis to allow for our lagged exposure (see below). PWH contributed person-time of observation from six months after cohort entry until cancer diagnosis, death, last laboratory value if >18 months before cohort data closure, or date of validated cancer data closure. PWH with extended periods out of care (defined as >18 months without laboratory values) were allowed to re-enter the observation period.

Our primary outcomes were incident invasive cancers, excluding basal and squamous cell skin cancers, which are not routinely captured in cancer registries and may not be completely ascertained in our data. We examined any incident cancer diagnosis as a composite outcome as well as individual AIDS-defining cancers (Kaposi sarcoma, non-Hodgkin lymphoma, and cervical cancer), HIV-associated cancers (Hodgkin lymphoma, liver cancer, anal cancer, head and neck cancer, and lung cancer), and the most common cancers in the general U.S. population (breast cancer, colorectal cancer, and prostate cancer). For those PWH with multiple cancers, only the first diagnosis of each cancer was included as an outcome.

We used time-updated, lagged values of CD4/CD8 ratio in our time-to-event analyses. We hypothesized a delayed effect of CD4/CD8 ratio and cancer diagnosis; therefore, we evaluated lagged CD4/CD8 ratio values, referencing the values recorded during defined intervals in the past in estimation of cancer risk during observation time (Supplemental Figure 1). Individual observation time was constructed based upon laboratory values and their corresponding dates of measurement. CD4 and CD8 cell counts were time-varying and carried forward for a maximum of one year. In the setting of a gap in laboratory values of more than one year, the remaining person-time was coded as missing laboratory values and the gap period was excluded from the analysis. Person-time of observation, including cancer diagnoses occurring during periods of missing laboratory values, were censored. Our primary analysis examined six-month lagged CD4/CD8 ratio value to minimize risk of reverse causality of CD4/CD8 ratio changes and cancer diagnosis. Given that the time between low CD4/CD8 ratio and cancer development is not known, we repeated analyses using 12-month, 18-month, and 24-month lagged CD4/CD8 ratio.

We used multivariable Cox proportional hazards models to calculate cause-specific hazard ratios for CD4/CD8 ratio and cancer risk [29]. Individuals were followed to their first cancer outcome or were censored if they died or had no event during their observed follow-up time.

Multivariable models included covariates selected *a priori* based on clinical knowledge and number of events (degrees of freedom) as potential confounders of the association between CD4/CD8 ratio and the cancer outcome of interest, other cancers, or death. All models included age at study entry, race/ethnicity (White, Black, Latinx or other), sex, year of cohort entry, history of HCV infection at any point during clinical follow-up, and time-varying status of history of any AIDS-defining illness (other than an ADC). Models examining risk factors for liver cancer additionally adjusted for history of hepatitis B virus (HBV) infection, history of injection drug use (IDU), history of heavy alcohol use (defined by documented diagnoses, medical record review, and survey results), and time-varying, lagged diagnosis of cirrhosis.

Time-varying lagged CD4 cell count and HIV RNA were included to examine the effect of CD4/CD8 ratio (independent of absolute CD4 cell count) and to account for the strong correlation between HIV viremia and the absolute CD4 and CD8 cell count. HIV RNA (log_10_ transformed), CD4 count (square root transformed), and CD4/CD8 ratio (natural logarithm transformed) were modeled as time-varying covariates. HIV RNA was used as a surrogate marker for time-varying ART use and values below than the limit of detection were calculated as (lower limit of detection - 1 copy/mL). To avoid assuming a linear relationship, most continuous covariates were included in the models using restricted cubic splines, including CD4/CD8 ratio values as there are no specific values associated with clinical outcomes in PWH [30]. For selection of consistent referent values for comparison of effects for each separate cancer analysis, we present adjusted hazard ratios (aHR) for the CD4/CD8 ratio comparing the 0.3 and 0.5 to 0.8 values which were selected for ease of comparison and approximation of the 25^th^, 50^th^, and 75^th^ percentile values of the six-month lagged CD4/CD8 ratio values for the study cohort. P-values were computed using Wald statistics. Models for sex-specific cancers (prostate and cervical) included only PWH of the relevant sex; breast cancer analyses included only females. Inclusion of all pre-specified covariates was dependent upon the number of cancer endpoints and reduced for breast and cervical cancer to avoid overfitting models. We stratified all Cox analyses by cohort to allow for separate baseline hazards.

In a sensitivity analysis, we fit models that included HIV acquisition risk factor; smoking status (ever vs. never); and six-month lagged, time-varying body mass index (BMI). When smoking status or alcohol was missing for individual patients, we used covariates and the outcome data to multiply impute (20 times) using predictive mean matching. Estimates from the imputation-specific analyses were then combined using Rubin’s rule [31].

All analyses were conducted in R version 3.6.2 and analysis code is available online at https://biostat.app.vumc.org/wiki/Main/ArchivedAnalyses. All *p* values were two-sided. NA-ACCORD data are publicly available for collaborative use, online at https://naaccord.org/.

## Results

The creation of the study population from the 15 eligible NA-ACCORD cohorts with contributing data is shown in Supplemental Figure 2. Characteristics of PWH included in the primary and sensitivity analyses are shown in Table 1. In the primary analysis using six-month lagged laboratory data, 83,893 PWH contributed 744,415 person-years of observation and 5,628 incident cancer diagnoses (excluding non-melanoma skin cancers). Of all observation time, 167,816 person-years (22.5%) were censored for missing CD4/CD8 ratio or HIV RNA levels. The median six-month lagged CD4/CD8 ratio was 0.52 (interquartile range: 0.30-0.82). PWH in both the primary and sensitivity analyses were predominantly male and less than half were white race. The median CD4 cell count, HIV RNA, and history of AIDS-defining illness at the start of observation time was similar between PWH included in the primary and sensitivity analyses. Table 1 also shows the frequency of the most common cancers diagnosed. For the primary analyses, there were a total of 5,628 incident cancers, of which 1,045 (19%) were an ADC. Of the NADCs, prostate cancer (n=817), lung cancer (n=755) and anal cancer (n=439) were the most frequent.

**Table 1.** Baseline patient characteristics and outcomes of PWH included in primary and sensitivity analyses of six-month-lagged laboratory data

|  | Primary analysis | Sensitivity analysis |
| --- | --- | --- |
|  | Total N = 83,893 | Total N = 40,975 |
|  | N (%) or median (IQR) | N (%) or median (IQR) |
| Sex |  |  |
| Male | 72,709 (87) | 32,420 (79) |
| Female | 11,184 (13) | 8,555 (21) |
| Age at study entry (years) | 43 (35-50) | 40 (33-47) |
| Race/ethnicity |  |  |
| White | 36,294 (43) | 19,648 (48) |
| Black | 31,252 (37) | 12,283 (30) |
| Latinx | 9,449 (11) | 5,728 (14) |
| Other or unknown | 6,898 (8) | 3,316 (8) |
| HIV transmission risk factor |  |  |
| Men who have sex with men | 28,725 (34) | 24,368 (59) |
| Injection drug use | 16,264 (19) | 4,200 (10) |
| Heterosexual | 10,927 (13) | 9,222 (23) |
| Other | 2,015 (2) | 1,007 (2) |
| Unknown or missing | 25,962 (31) | 2,178 (5) |
| Chronic hepatitis C virus infection | 17,855 (21) | 6,816 (16) |
| Chronic hepatitis B virus infection | 5,991 (7) | 3,023 (7) |
| Any history of heavy alcohol use<sup>a</sup> | 24,623 (29) | 11,500 (28) |
| Any history of tobacco use<sup>b</sup> | 28,373 (34) | 24,201(59) |
| CD4 cell count, cells/uL<sup>c</sup> | 413 (243-610) | 433 (262-631) |
| CD4/CD8 ratio<sup>d</sup> | 0.47 (0.27-0.76) | 0.50 (0.29-0.78) |
| Log<sub>10</sub> HIV RNA, copies/mL<sup>e</sup> | 2.3 (1.7-3.9) | 2.1 (1.7-3.8) |
| HIV RNA, copies/mL<sup>e</sup> |  |  |
| <500 | 41,470 (58) | 23,670 (60) |
| 500-4999 | 9807 (14) | 5,187 (13) |
| ≥5000 | 19,844 (28) | 10,784 (27) |
| Body mass index, kg/m<sup>2</sup> <sup>f</sup> | 25.2 (22.7-28.5) | 25.2 (22.6-28.5) |
| History of AIDS-defining illness | 11,438 (14) | 7,035 (17) |
| Geographic region of clinical care<sup>g</sup> |  |  |
| US Northeast | 10,788 (13) | 3,632 (9) |
| US Midwest | 7,403 (9) | 2,993 (7) |
| US Southeast | 20,483 (24) | 7,283 (18) |
| US West | 28,363 (33) | 19,209 (47) |
| US Southwest | 4,444 (5) | 111(0) |
| US Puerto Rico & Virgin Islands | 585 (1) | 2 (0) |
| Canada | 6,733 (8) | 3,293 (8) |
| Other | 12 (0) | 2 (0) |
| Unknown | 5,082 (6) | 4,450 (11) |
| Duration of follow-up, years | 8.5 (4.0-13.8) | 7.7 (3.6-12.7) |
| Initiated ART before study exit<sup>h</sup> | 75,201 (90) | 37,029 (90) |
| Proportion of follow-up time after ART |  |  |
| initiation |  |  |
| <10% | 834 (1) | 388 (1) |
| 10-24% | 895 (1) | 382 (1) |
| 25-49% | 2,171 (3) | 857 (2) |
| 50-74% | 4,108 (5) | 1,868 (5) |
| 75-89% | 4,042 (5) | 1,935 (5) |
| ≥ 90% | 63,528 (75) | 31,806 (78) |
| Missing ART data | 8,315 (10) | 3,739 (9) |
| Died | 14,345 (17) | 4,062 (10) |
| Any incident cancer<sup>i</sup> | 5,628 (7) | 2,172 (5) |
| AIDS-defining cancers |  |  |
| Non-Hodgkin lymphoma | 497 | 251 |
| Kaposi sarcoma | 501 | 253 |
| Cervix | 47 | 41 |
| HIV-associated cancers |  |  |
| Lung | 755 | 186 |
| Anus | 439 | 209 |
| Liver | 347 | 74 |
| Hodgkin lymphoma | 176 | 84 |
| Head and neck | 134 | 60 |
| Other |  |  |
| Prostate | 817 | 163 |
| Colorectum | 221 | 69 |
| Breast | 79 | 61 |
| Other | 1,615 | 721 |

IQR: interquartile range
<sup>a</sup>Ever documentation of alcohol abuse or at risk alcohol use. 2,165 (3%) missing in primary analysis, 80 (0%) missing in sensitivity analysis.
<sup>b</sup>Ever documentation of tobacco usage. 43,253 (52%) missing in primary analysis, 5,660 (14%) missing in sensitivity analysis.
<sup>c</sup>CD4 cell count at baseline (closest values within six months before or after). 13,170 (16%) missing in primary analysis, 1,197 (3%) missing in sensitivity analysis.
<sup>d</sup>CD8 and CD4/CD8 ratio at baseline (closest values within six months before or after). 15,915 (19%) missing in primary analysis, 2,198 (5%) missing in sensitivity analysis.
<sup>e</sup>HIV RNA at baseline (closest values with six months before or 30 days after). 12,772 (15%) missing in primary analysis, 1,334 (3%) missing in sensitivity analysis.
<sup>f</sup>Body mass index at baseline (closest values within six months before or after). 21,989 (26%) missing in primary analysis, 3,994 (10%) missing in sensitivity analysis.
<sup>g</sup> Geographic regions refers to location recorded closest to date of cohort entry. Other locations include Armed Forces Africa, Europe, Middle East, and Pacific and Guam.
<sup>h</sup> Study exit includes earliest occurrence of first cancer, death, or administrative censoring. ART data missing for 8,315 (9.9%) in primary analysis and 3,739 (9%) in sensitivity analysis.
<sup>i</sup> Any incident cancer excludes non-melanoma skin cancers.

Results of our primary analysis including the adjusted hazard ratios (aHR) and 95% confidence intervals (CI) comparing the 0.3 and 0.5 CD4/CD8 ratio values with the 0.8 are shown in Figure 1. There was a 24% increased risk of any incident cancer for individuals with six-month lagged CD4/CD8 ratio of 0.3 compared to 0.8, independent of CD4 cell count, HIV RNA value, and other covariates (aHR= 1.24, 95% CI 1.14-1.35). We observed the same pattern of increasing cancer risk with decreasing CD4/CD8 ratio for NHL, Kaposi sarcoma, lung cancer, anal cancer, and colorectal cancer. Six-month-lagged CD4/CD8 ratio was not associated with risk of cervical cancer, Hodgkin lymphoma, head and neck cancer, prostate cancer, breast cancer, nor liver cancer.

**Figure 1.**
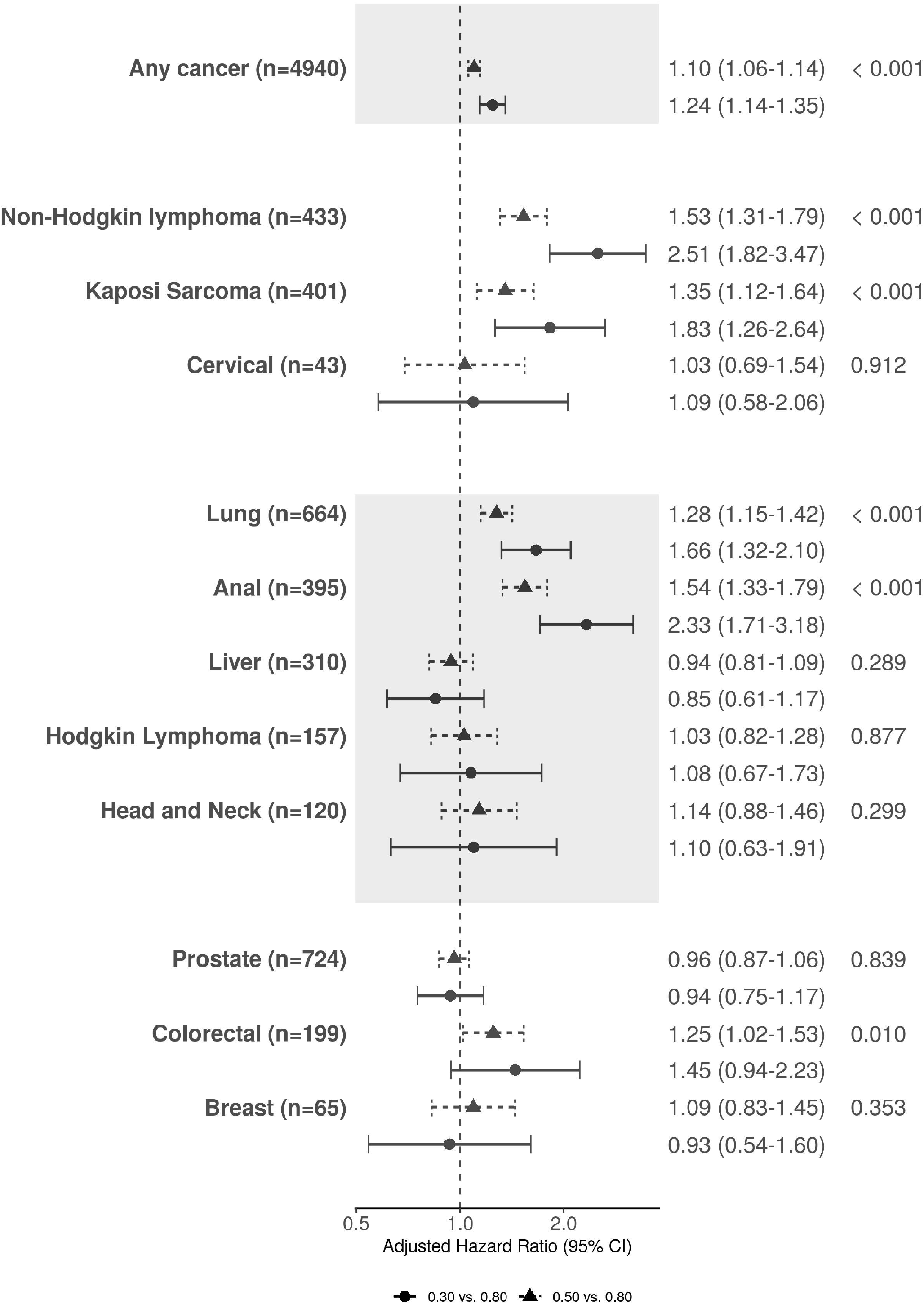
Adjusted hazard ratios and 95% confidence interval for cancer outcomes comparing the 6-month lagged, time-varying CD4/CD8 ratio values. Legend: Models for any cancer, lung cancer, Non-Hodgkin lymphoma, Hodgkin lymphoma, Kaposi sarcoma cancer, anal cancer, head and neck cancer, colorectal cancer included the covariates of age, sex, race, year of cohort entry, any history of chronic hepatitis C virus infection, and time-varying and time-updated CD4/CD8 ratio, CD4 cell count, HIV RNA, and history of AIDS-defining illness. The multivariable model for liver cancer included the covariates of age, sex, race, year of cohort entry, any history of chronic hepatitis C virus infection, history of chronic hepatitis B virus infection, history of heavy alcohol use, history of injection drug use, and time-varying and time-updated CD4/CD8 ratio, CD4 cell count, HIV RNA, history of AIDS-defining illness, and history of cirrhosis. The multivariable model for prostate cancer included only males and covariates age, race, year of cohort entry, any history of chronic hepatitis C virus infection, and time-varying and time-updated CD4/CD8 ratio, CD4 cell count, HIV RNA, and history of AIDS-defining illness. Models for breast cancer and cervical cancer included only females and included age, race (white vs. non-white), and time-varying and lagged CD4/CD8 ratio, CD4 cell count, HIV RNA, and history of AIDS-defining illness.

Results examining CD4/CD8 ratio values lagged at 12-months, 18-months, and 24-months as well as the six-months used in the primary analysis are shown in Supplemental Table 1.

Adjusted hazard ratios for cancers common among PWH along a continuum of CD4/CD8 values compared to 0.8 and using different lags are shown in Figure 2. Overall, we observed differing patterns of timing of low CD4/CD8 ratio and cancer risk across cancer types. As shown in Figure 2, low CD4/CD8 ratio values from six- to 24-months prior were all associated with increased risk of any cancer (*p* <0.001 for all time points). Similarly, low CD4/CD8 ratio was strongly associated with increased risk of anal and lung cancers across all lagged time points (*p* <0.001 for all time points for both cancers) (Figure 2). In contrast, while statistically significant at all lagged time points, low CD4/CD8 ratio was associated with highest risk of NHL in the six months prior, compared to 24 months (Figure 2d, *p*<0.01 for all lagged time points). In contrast, low CD4/CD8 ratio six months prior was not associated with risk of Hodgkin lymphoma (*p* =0.88) but was associated with increased risk 12 and 18-months prior (*p* = 0.052 and 0.015, respectively) (Figure 2). Additional figures for specific cancer types and lagged CD4/CD8 ratio values are shown in Supplemental Figure 3.

**Figure 2.**
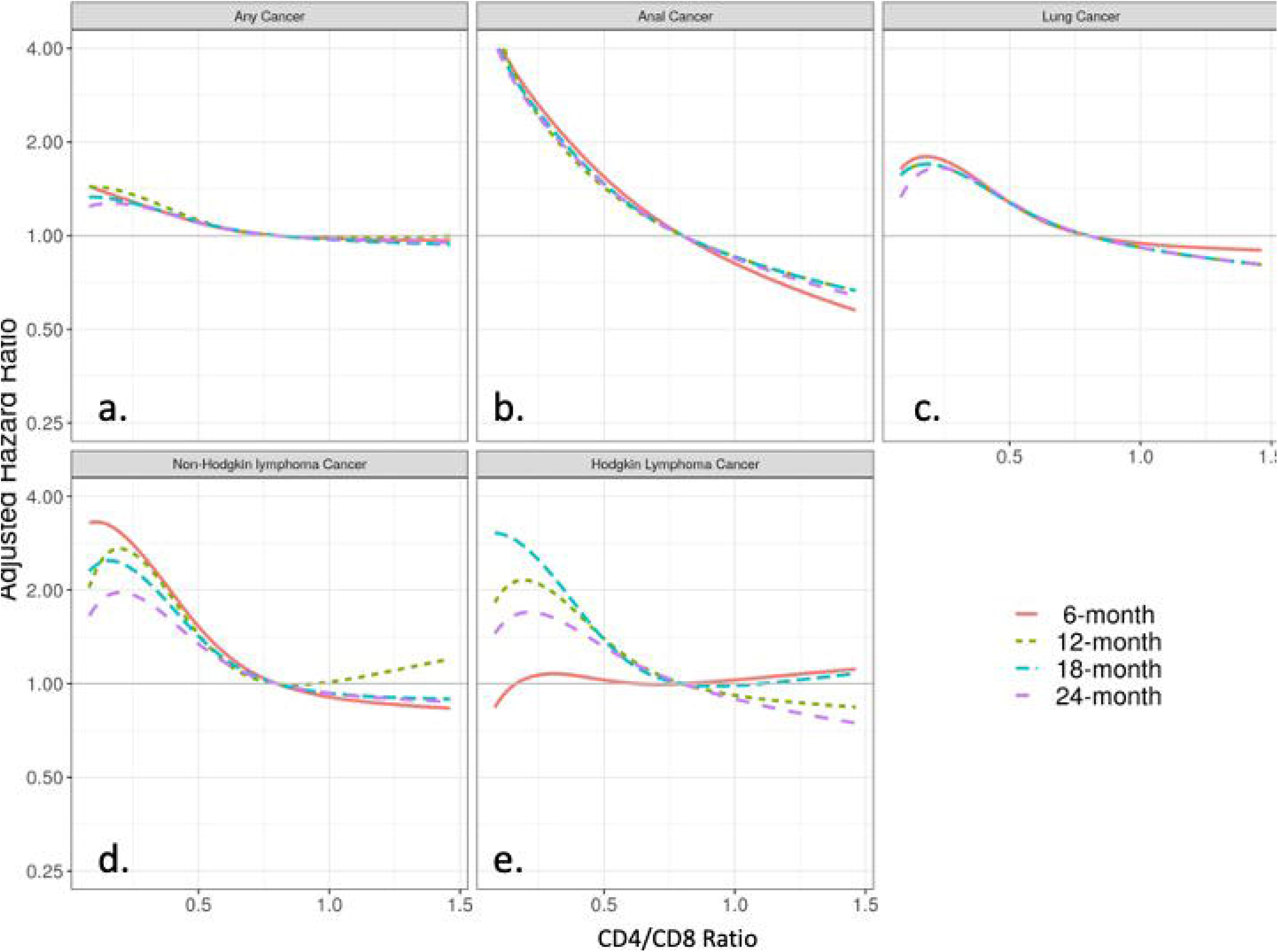
Adjusted hazard ratio for cancer and CD4/CD8 ratios lagged six, twelve, 18, and 24-months. Legend: Adjusted hazard ratio for CD4/CD8 ratio values lagged six months (solid orange lines), 12 months (dashed green lines), 18 months (dashed teal lines), and 24 months (dashed purple lines) for the following cancers (number of events for each model): (a) Any cancer (six months n=4,940; 12 months n=4,597; 18 months n=4,308; 24 months n=4,043); (b) Anal cancer (six months n=395; 12 months n=375; 18 months n=361; 24 months n=340); (c) Lung cancer (six months n=664; 12 months n=645; 18 months n=645; 24 months n=564); (d) NHL (six months n=433; 12 months n=382; 18 months n=333; 24 months n=310); (e) Hodgkin lymphoma (six months n=157; 12 months n=145; 18 months n=143; 24 months n=124). Models included the covariates of age, year of cohort entry, sex, race, any history of chronic hepatitis C virus infection, and time-varying and lagged CD4/CD8 ratio, CD4 cell count, HIV RNA, and history of AIDS-defining illness.

The multivariable analyses which included the subset of patients with available smoking, alcohol, HIV acquisition risk factor, and BMI data that assessed six-month lagged CD4/CD8 ratio and cancer risk are shown in Figure 3. Given the smaller number of cancers included in the sensitivity analyses, only any cancer, NHL, Kaposi sarcoma, lung cancer, and anal cancer were assessed in multivariable models to avoid over-fitting. The inverse association between low CD4/CD8 ratio and increased cancer risk persisted for any cancer, NHL, lung cancer, and anal cancer but was attenuated and no longer statistically significant for Kaposi sarcoma.

**Figure 3.**
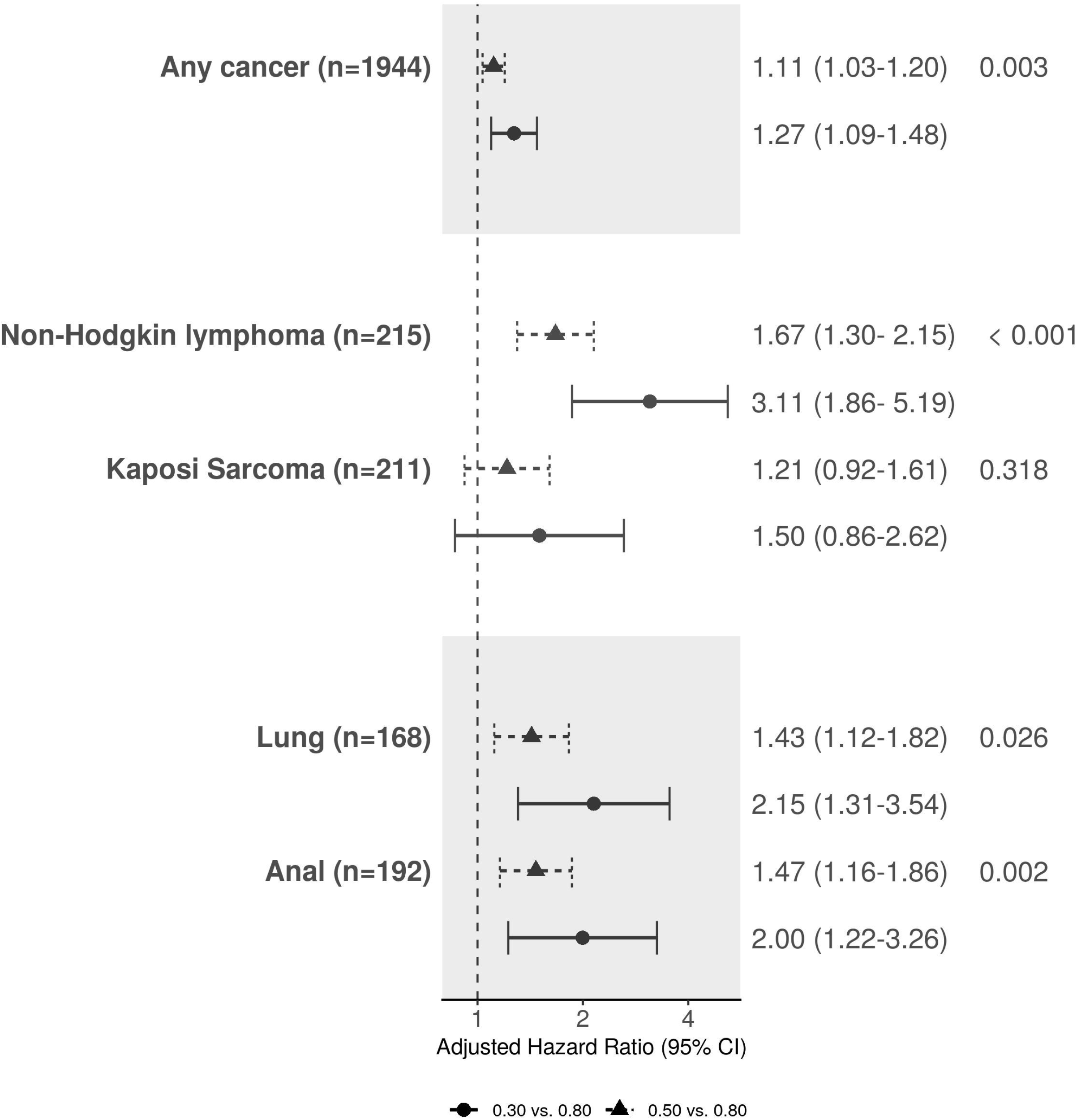
Sensitivity analyses for the adjusted hazard ratios and 95% confidence interval for cancer outcomes comparing the 6-month lagged, time-varying CD4/CD8 ratio values. Legend: Only models for any cancer, lung cancer, Non-Hodgkin lymphoma, Kaposi sarcoma cancer, and anal cancer performed due to number of events. All models included the covariates of age, sex, race, year of cohort entry, any history of chronic hepatitis C virus infection, history of smoking, history of heavy alcohol use, HIV acquisition risk factor, and time-varying and time-updated CD4/CD8 ratio, CD4 cell count, HIV RNA, history of AIDS-defining illness, and BMI.

## Discussion

In this large study of PWH in care and with high uptake of ART in the United States and Canada between 1998-2016, we found that low, time-lagged CD4/CD8 ratio was associated with increased risk of incident cancer diagnoses after accounting for CD4 cell count, HIV viral load, HIV acquisition risk, smoking, and other factors. The inverse association between CD4/CD8 ratio and cancer risk was observed for a number of specific cancers, including three of the most common cancers among PWH in the U.S.: NHL, lung cancer, and anal cancer. The association between low CD4/CD8 ratio and risk of cancer was not uniform, nor was the association exclusive for cancers associated with oncogenic viruses. Given the robust association up to 24-months prior to cancer diagnosis, CD4/CD8 ratio may indicate underlying immune dysfunction affecting carcinogenesis and inform cancer screening and prevention interventions among PWH.

Low CD4/CD8 ratio predicted risk of any cancer, independent of age, sex, CD4 cell count, HIV viral load, chronic HCV infection, and smoking history measures. Among PWH, low CD4/CD8 ratio is associated with older age; low CD4 nadir; male sex; HIV acquisition risk factors (particularly for MSM); chronic viral co-infection (including HCV and cytomegalovirus); and large HIV reservoir [27, 32-35]. Despite immune reconstitution from effective ART, PWH with low CD4/CD8 ratio have increased measures of adaptive immunosenescence including skewed T-lymphocytes populations from naïve to terminally differentiated phenotypes, higher expression of cellular markers of CD8 T cell activation and senescence, and higher kynurenine/trypotphan ratio [18, 20]. Low CD4/CD8 ratio has also been associated with increased measures of monocyte activation and systemic inflammatory measures, including IL-6, CRP, and soluble CD14 [18, 19]. Adaptive immune senescence, T cell activation, and inflammation have been identified as drivers of the excess risk of non-communicable diseases observed in PWH, including cardiovascular disease and cancers [36-39]. In PWH, low CD4/CD8 ratio has been associated with increased risk of mortality and non-communicable diseases [22, 23, 40]. Low CD4/CD8 has been associated with risk of NADCs in smaller studies of PWH [24, 41]. To our knowledge, this is the largest study to demonstrate an independent association between low CD4/CD8 ratio and cancer risk in PWH, which we found was consistent up to two-years preceding the cancer diagnosis.

We found that NHL, lung cancer and anal cancer, three of the leading causes of cancer-related morbidity and mortality in PWH in the U.S., were strongly and consistently associated with low CD4/CD8 ratio [42]. Both lung cancer and anal cancer have higher incidence and are diagnosed at younger ages in PWH compared to the general population, suggesting accelerated aging biology, such as immunosenescence, may contribute to cancer risk in PWH [6, 8, 43, 44]. The large Veterans Aging Cohort Study found an independent association between low, cumulatively-averaged CD4/CD8 ratio and lung cancer risk in U.S veterans with HIV [25]. Our results are consistent with their findings; supporting the hypothesis that immune dysfunction, independent of oncogenic viral activity, may contribute to cancer risk in PWH.

This is the largest study to show low peripheral CD4/CD8 ratio predicts anal cancer risk, independent of HIV acquisition risk factor, smoking, or immunodeficiency. A single-center study of PWH observed that low CD4/CD8 ratio nadir and CD4/CD8 proximal to anal cancer screening were associated with high-grade anal dysplasia and cancer; however, analyses did not adjust for concurrent CD4 cell count [45]. The ratio of CD4/CD8 tumor-infiltrating and stromal lymphocytes in tissue biopsies of HPV-associated pre-cancers of the cervix, anus, and head and neck have been associated with disease progression or regression; however, circulating CD4/CD8 ratio has not been evaluated in anal pre-cancers [46-49]. ART has not been shown to be associated with the regression or clearance of anal lesions, suggesting immune restoration does not entirely eliminate the increased risk [50, 51]. That we did not find an association between CD4/CD8 ratio and cervical cancer may be due to the small number of cancer cases in this cohort. Additionally, we did not find a significant association between CD4/CD8 ratio and head and neck cancer; however, we could not distinguish cases that were associated with HPV. Further research is needed to assess CD4/CD8 ratio and HPV-associated and HPV-negative head and neck cancers.

There are important limitations of this study. The study population was disproportionately male, not only resulting in low numbers of cervical cancer and breast cancer but limiting our ability to examine CD4/CD8 ratio and cancer risk in women with HIV more generally. HIV acquisition risk factor and behavioral data including smoking history were not available from all cohorts, though results from our sensitivity analyses were generally consistent with those from the primary analyses. However, for lung cancer in particular, our results may still be affected by residual confounding from smoking given the limited smoking information in our cohort.

CD4/CD8 ratio is lower in settings of chronic viral infections and our data on prevalent oncogenic viruses were limited to HCV and HBV. The cancer most associated with these viruses, liver cancer, was not significantly associated with CD4/CD8 ratio in multivariable models. Seroprevalence of CMV, human herpes virus 8 (Kaposi sarcoma), Epstein-Barr virus (Hodgkin lymphoma and NHL), or infection with high-risk HPV (cervical, anal, and head and neck cancers) is not universally collected in clinical care of PWH. Our data may also be subject to misclassification bias as a result of carrying-forward CD4/CD8 data and periods of missing CD4/CD8 data.

In the largest study of PWH in the U.S. and Canada, we found that lagged, low CD4/CD8 ratio was associated with increased risk of certain incident cancers. Low CD4/CD8 ratio was associated with increased risk of NHL, lung cancer, and anal cancer. The consistency of association observed up to 24-months prior to cancer diagnosis suggests CD4/CD8 ratio may be a useful biomarker for screening for lung and anal cancer in PWH where the average age at diagnosis is younger than in the general population. Further investigation into the roles of immunosenescence, immune activation, and inflammation and cancer risk observed in PWH is needed.

## Data Availability

NA-ACCORD welcomes interested investigators to collaborate with us for use of our data. Please visit https://naaccord.org/ for additional information.

## NA-ACCORD Collaborating Cohorts and Representatives

**AIDS Clinical Trials Group Longitudinal Linked Randomized Trials**: Constance A. Benson and Ronald J. Bosch

**AIDS Link to the IntraVenous Experience**: Gregory D. Kirk

**Emory-Grady HIV Clinical Cohort**: Vincent Marconi and Jonathan Colasanti

**Fenway Health HIV Cohort**: Kenneth H. Mayer and Chris Grasso

**HAART Observational Medical Evaluation and Research**: Robert S. Hogg, Viviane Lima, P. Richard Harrigan, Julio SG Montaner, Benita Yip, Julia Zhu, and Kate Salters

**HIV Outpatient Study**: Kate Buchacz and Jun Li

**HIV Research Network**: Kelly A. Gebo and Richard D. Moore

**Johns Hopkins HIV Clinical Cohort**: Richard D. Moore

**John T. Carey Special Immunology Unit Patient Care and Research Database, Case Western Reserve University**: Jeffrey M. Jacobson

**Kaiser Permanente Mid-Atlantic States**: Michael A. Horberg

**Kaiser Permanente Northern California**: Michael J. Silverberg

**Longitudinal Study of Ocular Complications of AIDS**: Jennifer E. Thorne

**MACS/WIHS Combined Cohort Study**: Todd Brown, Phyllis Tien, and Gypsyamber D’Souza

**Maple Leaf Medical Clinic**: Graham Smith, Mona Loutfy, and Meenakshi Gupta

**The McGill University Health Centre, Chronic Viral Illness Service Cohort**: Marina B. Klein

**Multicenter Hemophilia Cohort Study–II**: Charles Rabkin

**Ontario HIV Treatment Network Cohort Study**: Abigail Kroch, Ann Burchell, Adrian Betts, and Joanne Lindsay

**Parkland/UT Southwestern Cohort**: Ank Nijhawan

**Retrovirus Research Center, Universidad Central del Caribe, Bayamon Puerto Rico**: Angel M. Mayor

**Southern Alberta Clinic Cohort**: M. John Gill

**Study of the Consequences of the Protease Inhibitor Era**: Jeffrey N. Martin

**Study to Understand the Natural History of HIV/AIDS in the Era of Effective Therapy**: Jun Li and John T. Brooks

**University of Alabama at Birmingham 1917 Clinic Cohort**: Michael S. Saag, Michael J. Mugavero, and James Willig

**University of California at San Diego**: Laura Bamford and Maile Karris

**University of North Carolina at Chapel Hill HIV Clinic Cohort**: Joseph J. Eron and Sonia Napravnik

**University of Washington HIV Cohort**: Mari M. Kitahata and Heidi M. Crane

**Vanderbilt Comprehensive Care Clinic HIV Cohort**: Timothy R. Sterling, David Haas, Peter Rebeiro, and Megan Turner

**Veterans Aging Cohort Study**: Lesley Park and Amy Justice

**NA-ACCORD Study Administration:**

**Executive Committee:** Richard D. Moore, Keri N. Althoff, Stephen J. Gange, Mari M. Kitahata, Jennifer S. Lee, Michael S. Saag, Michael A. Horberg, Marina B. Klein, Rosemary G. McKaig, and Aimee M. Freeman

**Administrative Core:** Richard D. Moore, Keri N. Althoff, and Aimee M. Freeman

**Data Management Core:** Mari M. Kitahata, Stephen E. Van Rompaey, Heidi M. Crane, Liz Morton, Justin McReynolds, and William B. Lober

**Epidemiology and Biostatistics Core:** Stephen J. Gange, Jennifer S. Lee, Brenna Hogan, Bin You, Elizabeth Humes, Lucas Gerace, Cameron Stewart, and Sally Coburn

