## Supplemental Material for "Low CD4/CD8 ratio predicts cancer risk among adults with HIV"

Supplemental Figure 1. Conceptual diagram of time-varying and time-fixed covariates used in multivariable, time-to-event analyses

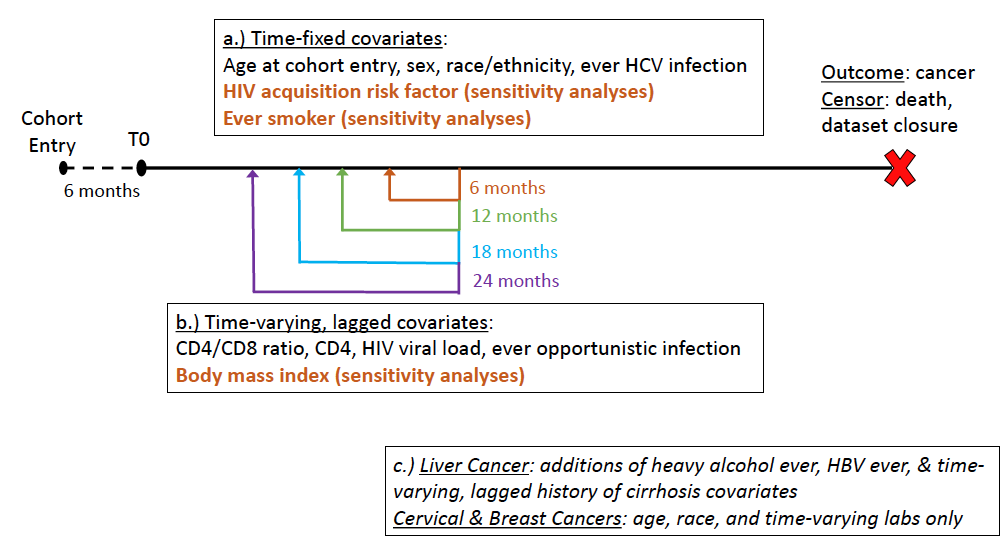

Legend: Black line indicates observation time from T0 to cancer event or death. Box (a) includes list of time-fixed covariates included in all models (black) and additional covariates included in sensitivity analyses (orange, bold). Box (b) lists the time-varying covariates included in all models (black) and added in sensitivity models (orange, bold). Time-varying covariates were lagged such that values referenced at a given point in during observation corresponded to recorded laboratory values or prevalent diagnoses (in the case of opportunistic infections) six (orange), twelve (green), eighteen (blue), and twenty-four (purple) months prior. Box (c) indicates additional time-fixed and time-varying covariates included in models for liver cancer and restricted list of covariates included in models for cervical and breast cancers.

Supplemental Figure 2. Study population flow chart

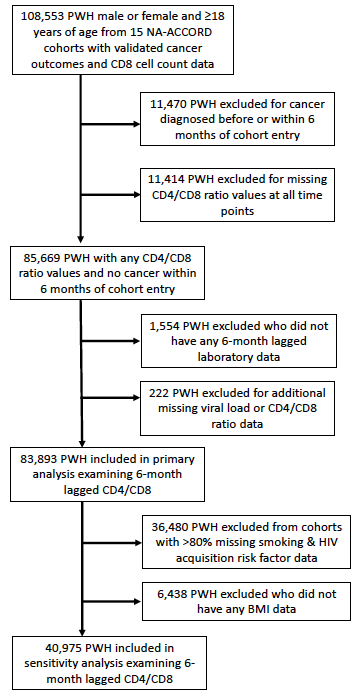

Supplemental Figure 3. Adjusted hazard ratio for cancer and CD4/CD8 ratios lagged six, twelve, 18, and 24-months.

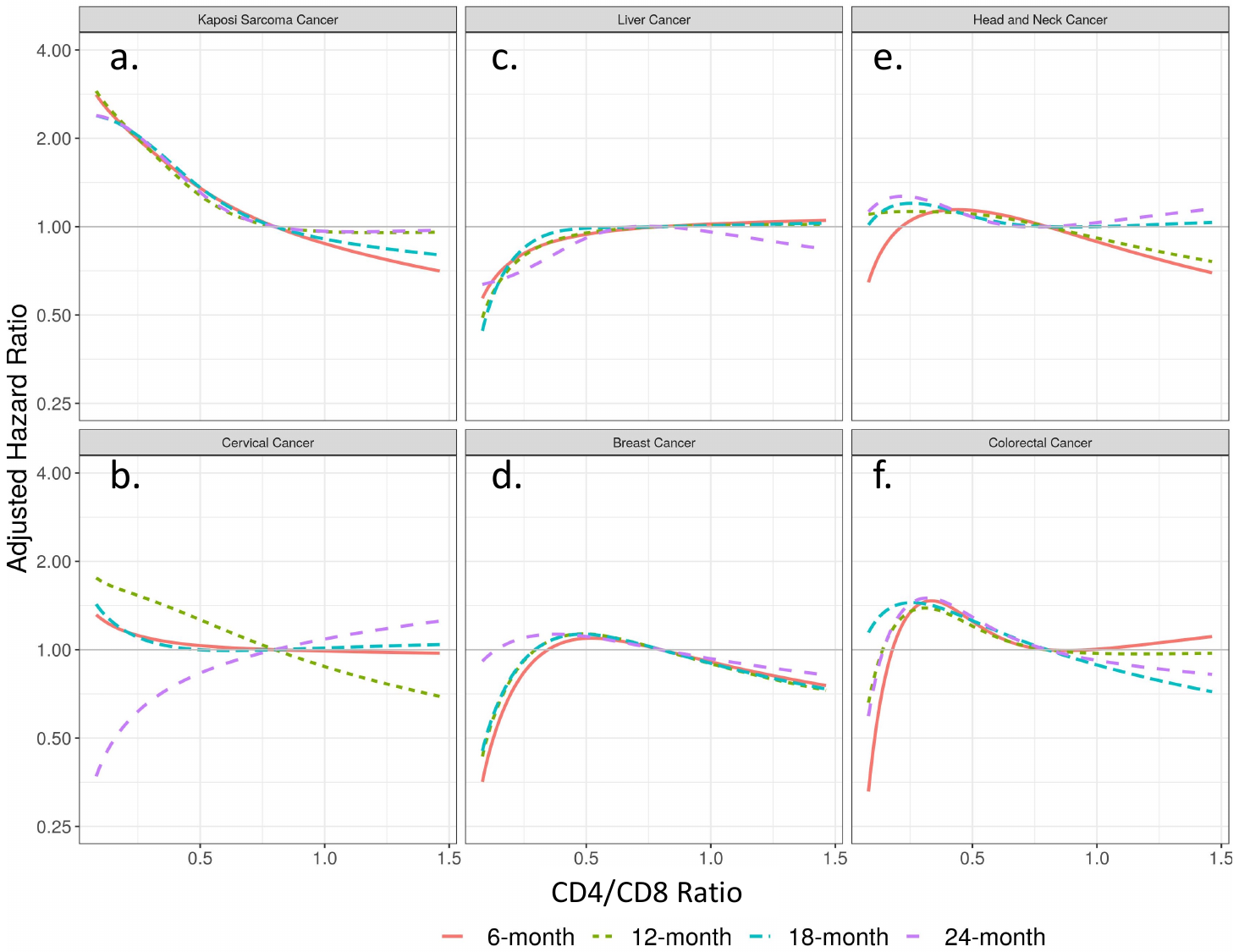

Legend: Adjusted hazard ratio for CD4/CD8 ratio values lagged six months (solid orange lines), 12 months (dashed green lines), 18 months (dashed teal lines), and 24 months (dashed purple lines) for the following cancers (number of events for each model): (a) Kaposi sarcoma (six months n=401; 12 months n=346; 18 months n=298; 24 months n=289); (b) Cervical cancer (six months n=43; 12 months n=42; 18 months n=31; 24 months n=27); (c) Liver cancer (six months n=310; 12 months n=286; 18 months n=286; 24 months n=268); (d) Breast cancer (six months n=65; 12 months n=62; 18 months n=54; 24 months n=50); (e) Head and neck cancer (six months n=120; 12 months n=117; 18 months n=113; 24 months n=105); (f) Colorectal cancer (six months n=199; 12 months n=186; 18 months n=181; 24 months n=171). Models for Kaposi sarcoma, liver cancer, and head and neck cancer included the covariates of age, sex, race, year of cohort entry, any history of chronic hepatitis C virus infection, and time-varying and lagged CD4/CD8 ratio, CD4 cell count, HIV RNA, and history of AIDS-defining illness. Liver cancer models additionally included history of hepatitis B virus, any history of heavy alcohol use, history of injection drug use, and time-updated, lagged history of cirrhosis. Models for breast cancer and cervical cancer restricted to females only and included age, race (white vs. non-white), and time-varying and lagged CD4/CD8 ratio, CD4 cell count, HIV RNA, and history of AIDS-defining illness only due to small sample size.

Supplemental Table 1. The adjusted association between CD4/CD8 ratio and time to first cancer events

|  | | **Lagged 6 months** | **Lagged 12 months** | **Lagged 18 months** | **Lagged 24 months** |
| --- | --- | --- | --- | --- | --- |
| **Cancer event** | **CD4/CD8** | **HR (95%CI)** | **HR (95%CI)** | **HR (95%CI)** | **HR (95%CI)** |
| **Any Cancer** | 0.30 vs. 0.80 | 1.24 (1.14-1.35) | 1.31 (1.20-1.43) | 1.24 (1.13-1.36) | 1.23 (1.12-1.35) |
|  | 0.50 vs. 0.80 | 1.10 (1.06-1.14) | 1.12 (1.08-1.17) | 1.11 (1.06-1.15) | 1.10 (1.06-1.15) |
| **Non-Hodgkin lymphoma** | 0.30 vs. 0.80 | 2.51 (1.82-3.47) | 2.42 (1.72-3.39) | 2.15 (1.50-3.09) | 1.84 (1.27-2.66) |
|  | 0.50 vs. 0.80 | 1.53 (1.31-1.79) | 1.44 (1.23-1.69) | 1.42 (1.19-1.70) | 1.34 (1.12-1.61) |
| **Kaposi Sarcoma** | 0.30 vs. 0.80 | 1.83 (1.26-2.64) | 1.81 (1.25-2.63) | 1.89 (1.27-2.82) | 1.88 (1.27-2.79) |
|  | 0.50 vs. 0.80 | 1.35 (1.12-1.64) | 1.28 (1.06-1.54) | 1.36 (1.11-1.66) | 1.31 (1.08-1.59) |
| **Cervical Cancer** | 0.30 vs. 0.80 | 1.09 (0.58-2.06) | 1.48 (0.74-2.96) | 1.06 (0.50-2.26) | 0.67 (0.35-1.29) |
|  | 0.50 vs. 0.80 | 1.03 (0.69-1.54) | 1.27 (0.81-1.97) | 1.00 (0.62-1.61) | 0.83 (0.55-1.25) |
| **Lung Cancer** | 0.30 vs. 0.80 | 1.66 (1.32-2.10) | 1.60 (1.26-2.02) | 1.6 (1.26-2.02) | 1.61(1.25-2.07) |
|  | 0.50 vs. 0.80 | 1.28 (1.15-1.42) | 1.28 (1.15-1.42) | 1.28 (1.15-1.42) | 1.29 (1.15-1.45) |
| **Anal Cancer** | 0.30 vs. 0.80 | 2.33 (1.71-3.18) | 2.12 (1.55-2.91) | 2.21 (1.59-3.06) | 2.15 (1.53-3.02) |
|  | 0.50 vs. 0.80 | 1.54 (1.33-1.79) | 1.43 (1.23-1.66) | 1.47 (1.26-1.72) | 1.46 (1.24-1.72) |
| **Liver Cancer** | 0.30 vs. 0.80 | 0.85 (0.61-1.17) | 0.85 (0.60-1.19) | 0.90 (0.64-1.27) | 0.75 (0.52-1.06) |
|  | 0.50 vs. 0.80 | 0.94 (0.81-1.09) | 0.95 (0.82-1.11) | 0.99 (0.85-1.15) | 0.91 (0.78-1.07) |
| **Hodgkin Lymphoma** | 0.30 vs. 0.80 | 1.08 (0.67-1.73) | 1.98 (1.19-3.30) | 2.24 (1.33-3.76) | 1.64 (0.95-2.83) |
|  | 0.50 vs. 0.80 | 1.03 (0.82-1.28) | 1.40 (1.10-1.78) | 1.39 (1.09-1.76) | 1.31 (1.01-1.70) |
| **Head and Neck Cancer** | 0.30 vs. 0.80 | 1.10 (0.63-1.91) | 1.13 (0.64-1.98) | 1.19 (0.67-2.12) | 1.24 (0.69-2.24) |
|  | 0.50 vs. 0.80 | 1.14 (0.88-1.46) | 1.11 (0.86-1.42) | 1.08 (0.84-1.40) | 1.08 (0.83-1.40) |
| **Prostate Cancer** | 0.30 vs. 0.80 | 0.94 (0.75-1.17) | 1.02 (0.82-1.28) | 0.95 (0.75-1.20) | 0.96 (0.75-1.22) |
|  | 0.50 vs. 0.80 | 0.96 (0.87-1.06) | 0.98 (0.89-1.09) | 0.97 (0.87-1.08) | 0.96 (0.86-1.07) |
| **Colorectal Cancer** | 0.30 vs. 0.80 | 1.45 (0.93-2.27) | 1.41 (0.89-2.22) | 1.48 (0.93-2.35) | 1.52 (0.94-2.46) |
|  | 0.50 vs. 0.80 | 1.25 (1.02-1.53) | 1.21 (0.99-1.48) | 1.27 (1.03-1.57) | 1.30 (1.05-1.62) |
| **Breast Cancer** | 0.30 vs. 0.80 | 0.93 (0.54-1.60) | 1.01 (0.58-1.75) | 1.01 (0.57-1.79) | 1.12 (0.62-2.02) |
|  | 0.50 vs. 0.80 | 1.09 (0.83-1.45) | 1.13 (0.85-1.51) | 1.13 (0.82-1.56) | 1.11 (0.81-1.52) |

Adjusted Hazard Ratios and 95% confidence interval for different cancer were estimated using the multivariable Cox proportional hazards models. Models for any cancer, lung cancer, Non-Hodgkin lymphoma, Kaposi sarcoma cancer, anal cancer, head and neck cancer, colorectal cancer included the covariates of age, sex, race, any history of chronic hepatitis C virus infection, year of cohort entry, and time-varying and time-updated CD4/CD8 ratio, CD4 cell count, HIV RNA, and history of AIDS-defining illness. The multivariable model for liver cancer included the covariates of age, sex, race, any history of chronic hepatitis C virus infection, history of chronic hepatitis B virus infection, history of heavy alcohol use, history of injection drug use, year of cohort entry, and time-varying and time-updated CD4/CD8 ratio, CD4 cell count, HIV RNA, history of AIDS-defining illness, and history of cirrhosis. The multivariable model for prostate cancer included only males and covariates age, race, any history of chronic hepatitis C virus infection, and time-varying and time-updated CD4/CD8 ratio, CD4 cell count, HIV RNA, year of cohort entry, and history of AIDS-defining illness. Models for breast cancer and cervical cancer included only females and included age, race (white vs. non-white), and time-varying and time-updated CD4/CD8 ratio, CD4 cell count, HIV RNA, and history of AIDS-defining illness.

Abbreviations used:

HR: hazard ratio

CI: confidence interval
